# Facilitators for green behaviour change during GP consultations: a qualitative study

**DOI:** 10.64898/2026.11.25.26368954

**Authors:** E.H. Visser, S. Koivusaari, I.A. Slootweg, E.A. Brakema, H.M.M. Vos, M.A. Adriaanse

**Affiliations:** Health Campus The Hague, Department of Public Health and Primary Care, Leiden University Medical Centre; Department of Health, Medical and Neuropsychology of the Faculty of Social and Behavioural Sciences, Leiden University

**Author notes:** **Contact information** Correspondence may be addressed to Eva H. Visser, Department of Public Health and Primary Care, Leiden University Medical Center, Postbus 9600, 2300 RC Leiden, Netherlands;. Eva H. Visser, Health Campus The Hague, Department of Public Health and Primary Care, Leiden University Medical Centre, Netherlands. Satu Koivusaari, Department of Health, Medical and Neuropsychology of the Faculty of Social and Behavioural Sciences, Leiden University. Evelyn A. Brakema, Health Campus The Hague, Department of Public Health and Primary Care, Leiden University Medical Centre. Irene A. Slootweg, Health Campus The Hague, Department of Public Health and Primary Care, Leiden University Medical Centre. Hedwig M.M. Vos, Health Campus The Hague, Department of Public Health and Primary Care, Leiden University Medical Centre. Marieke A. Adriaanse, Health Campus The Hague, Department of Public Health and Primary Care, Leiden University Medical Centre, and Department of Health, Medical and Neuropsychology of the Faculty of Social and Behavioural Sciences of Leiden University.

**Keywords:** General Practice, Climate Change, Sustainable Development, Behaviour, Health Personnel, Qualitative Research

## Abstract

**Background:** GP consultations are a major source of primary care’s environmental impact and therefore a promising point of intervention for mitigation. Understanding how pioneers, green GPs, have successfully changed their behaviour during these consultations, may support other GPs to follow suit.

**Aim:** To identify key facilitators for GPs to change their behaviour from environmentally less sustainable to environmentally more sustainable during consultations.

**Design and setting:** In 2025, we conducted a qualitative study with Dutch green GPs, defined as GPs with knowledge of and interest in sustainable healthcare, and who apply sustainable behaviours during consultations.

**Method:** We used semi-structured interviews guided by three vignettes about green behaviour changes: sustainable prescribing of inhalation medication, prescribing less per consultation, and focussing on lifestyle changes before starting preventive cardiovascular medication. We deductively coded the facilitators using the Theoretical Domains Framework (TDF), followed by inductive coding using thematic analysis.

**Results:** We interviewed 12 green GPs and found that the key facilitators loaded on the TDF domains: knowledge (fostering moments of realisation), environmental context and resources (a supportive green workplace), social influence (a green network) and beliefs about consequences (focusing on benefits).

**Conclusion:** Changing GPs’ behaviour during consultations to be environmentally more sustainable begins with realising the impact of healthcare on the planet, and is sustained by focusing on the benefits of sustainable healthcare and creating opportunities within physical and social environments. Building on these facilitators, interventions should focus on altering care environments to enable GPs to easily opt for the sustainable option.

**How this fits in:** GP consultations are a major source of primary care’s environmental impact and therefore a promising point of intervention for mitigation, enabling individual GPs to directly improve the sustainability of care through changes in their behaviour during consultations. Green GPs, who have already implemented environmentally sustainable changes in their practices, demonstrate that such changes are possible. In this study we build upon these experiences and identified key facilitators for GPs to make a behaviour change from environmentally less sustainable to environmentally more sustainable during consultations. We found that the key facilitators of behaviour change include fostering a moment of realisation to start the behaviour change, and emphasising the benefits of sustainable healthcare for patients, society, GPs, and the planet, and designing physical and social environments that encourage its provision.

## Introduction

Accounting for more than 20% of healthcare-related carbon emissions,(1, 2) primary care carries responsibility, but also offers opportunity, for mitigating healthcare’s environmental impact. A considerable part of this impact originates in consultations, through prescriptions (i.e. pharmaceuticals) and ordering laboratory tests (i.e. chemicals).(1, 3) Therefore, consultations represent an area where individual general practitioners (GPs) can directly influence the environmental impact of care by changing their behaviour. A prototypical example of such a ‘green’ behaviour change is prescribing a more environmentally sustainable medication option, e.g. switching from a pressurised metered-dose inhaler (pMDI) to a dry-powder inhaler (DPI).(4)

As research into sustainable GP care grows,(5-8) an increasing number of such sustainable alternatives are being identified,(9, 10) and incorporated into clinical guidelines.(11) Yet simply having these alternatives available, even included in guidelines, is often not enough to change health professional behaviour.(12) Concerns about jeopardising the doctor-patient relationship when discussing sustainable care have been identified as a barrier by GPs.(13) However, the actual barrier may lie in GPs’ expectations, as research examining patients’ responses to sustainable alternatives has shown that patient satisfaction remains high when sustainable alternatives are proposed.(14) This discrepancy between the perceived barrier and patients’ actual responses highlights the complexity of behaviour change and underscores the fact that behaviour is influenced by a broad range of factors.(15) Understanding these factors could support GPs in changing their behaviour during consultations to become more environmentally sustainable.

To investigate this, it may be useful to adopt a behavioural science perspective. If we view sustainable GP care through this lens, we may be able to zoom out and uncover the underlying domains on which the behaviour change is built, such as making sustainability the norm or aligning the new behaviour with one’s professional identity. These domains are captured in the Theoretical Domains Framework (TDF) and the overarching COM-B model (Capability, Opportunity and Motivation interact to produce Behaviour);(16, 17) frameworks developed to identify influences on health professional behaviour. To understand which domains facilitate the adoption of sustainable GP care, it may be particularly informative to focus on GPs who already made this transition in practice, i.e. ‘green GPs’. As early adopters, they can offer valuable insights into the factors that helped them make and sustain green changes.

Therefore, in this study, we aim to identify key facilitators for GPs to change their behaviour from environmentally less sustainable to environmentally more sustainable during consultations. By focusing on what has worked for those who have already made this transition, we seek to generate insights that are both empirically grounded and practically applicable.

## Methods

We conducted a two-phase sequential qualitative study. We followed COREQ guidelines, where applicable, as detailed in Supplementary Table S1.(18) Phase one comprised semi-structured interviews to identify facilitators and phase two included focus groups to prioritize the found facilitators based on helpfulness and feasibility.

The study was approved by the non-WMO committee of the Leiden University Medical Centre (#25-3036). We collected data from April to June 2025. The protocol and logbook will be available on the Open Science Framework after publication. Raw data is not available to ensure privacy of our participants.

### Participants and recruitment

Our sample consisted of Dutch, fully qualified, green GPs. We defined a green GP as a GP with knowledge of and interest in sustainable healthcare, and who applies more sustainable behaviours during consultations. We recruited participants through purposive sampling (variation: gender, age, experience as a GP and with sustainable healthcare, and workplace type) and snowballing. We approached members of ‘de Groene Huisarts’ (‘The Dutch Green GP’), as well as other green GPs who opted to be contacted in the small pilot study, using a messaging platform (Signal Messenger LLC, Mountain View, CA, USA) or email. Data collection continued until theoretical saturation was reached.(19) We obtained digital informed consent prior to participation.

### Study design

In phase one EV performed online semi-structured interviews, using vignettes to elicit facilitators closely reflecting real-life behaviour change. The vignettes were: sustainable prescribing of inhalation medication, prescribing less per consultation, and focussing on lifestyle changes before starting preventive cardiovascular medication. The vignettes were based on a pilot questionnaire study among 24 green oriented Dutch GPs as detailed in Supplement 2 and written using Atkins et al.’s specificity rules.(16) The participant chose which vignette to start with. The interview guide and vignettes are available in Supplement 3.The interviews were held via Microsoft Teams (Microsoft Corporation, Redmond, WA, USA) and audio recorded.

In phase two we conducted online focus groups via Microsoft Teams (Microsoft Corporation, Redmond, WA, USA), moderated by IS and observed by EV; sessions were audio-recorded. Following the Prioritizing Implementation Barriers toolkit,(19) participants individually rated actionable facilitators for helpfulness and feasibility using a four-point Likert scale in WooClap (WooClap SA, Brussels, Belgium). Ratings were then displayed in a two-by-two matrix and discussed by the participants. The topic guide is available in Supplement 4.

The two-phased approach was intended to reduce an extensive list of facilitators to a smaller set of priorities. However, the interviews already identified key facilitators, and focus groups added little further prioritisation because participants rated the facilitators highly on both criteria. We therefore discuss phase two only briefly.

### Analysis

We transcribed the audio files verbatim using Microsoft Teams (Microsoft Corporation, Redmond, WA, USA), audio checked by EV. We analysed the data in Atlas.ti 9 (ATLAS.ti Scientific Software Development GmbH, Berlin, Germany). As a GP trainee familiar with medical jargon, EV reviewed all transcripts and coded text fragments describing potential facilitators. We analysed from a realist approach and used a hybrid deductive-inductive thematic analysis.(20) We first coded facilitators to the TDF,(16) with EV coding all transcripts and SK independently coding 50% of the data in three phases, after which coding decisions were compared and discrepancies resolved through discussion with MA.

EV and SK then collaboratively conducted inductive thematic analysis within each TDF domain using theme piles.(20) Key facilitators were identified based on their endorsement and perceived importance by participants. Of note, participants identified both facilitators and strategies for sustainable behaviours. As these were often inseparable, both were coded within the relevant TDF domain and are hereafter referred to as ‘facilitators’.

Overlapping themes were assigned to the best-fitting domain and cross-referenced. Before the in-depth analysis described above, we exploratively reviewed the interviews to generate a longlist of actionable facilitators that informed the focus groups. We analysed the focus groups using a two-by-two matrix and descriptive statistics.

### Reflexivity statement

The research team consisted of members with diverse backgrounds, including general practice, behavioural - and educational sciences. Throughout the study, we held regular meetings to discuss and reflect on decisions relating to the design, data collection, analysis, and drafting of the manuscript. The authors are to varying degrees, involved in promoting sustainability, or sustainable healthcare specifically. The investigator who conducted the interviews and focus groups (EV), was trained in qualitative interviewing, and knew several of the participants prior to their participation in this study. The research team’s involvement in sustainable healthcare facilitated good access to the target population. However, this also necessitated careful reflection on the balance between our roles as researchers and as fellow sustainable healthcare professionals. During the interviews, EV was at times able to manage this dual role effectively, while at other moments this proved more challenging.

Consequently, some interviews did not go into as much depth as would have been possible. On the other hand, this shared professional context appeared to help participants feel at ease from the outset and may have facilitated openness in sharing their experiences. We do acknowledge our team’s potential ‘green’ lens. This may have led to a more positive view of sustainable healthcare.(21)

## Results

Between April and June 2025, we conducted 12 interviews and two focus groups (n=4; n=5). Around 80% of participants were women. The interviews lasted on average 49 minutes [range 35 – 58]. The focus groups lasted 56 and 60 minutes, respectively. The participant characteristics are outlined in table 1.

**Table 1.** Characteristics of green GPs participating in the interviews and focus groups on green behaviour change in clinical consultations [Data are presented as % or mean (range)]

| Characteristic | Interviews (n=12) |  | Focus groups (n=9) |  |
|---|---|---|---|---|
| Gender, women | 83% (n=10) |  | 78% (n=7) |  |
| Age, years | 45 (34-63) |  | 47 (37-63) |  |
| Professional experience as GP, years | 12 (1-34) |  | 13 (2-34) |  |
| Professional experience with sustainable healthcare, years | 6 (3-15) |  | 7 (3-15) |  |
| GP role | GP partner | 42% (n=5) | GP partner | 33% (n=3) |
|  | Salaried GP | 8% (n=1) | Salaried GP | 11% (n=1) |
|  | Locum GP | 25% (n=3) | Locum GP | 22% (n=2) |
|  | Locum and salaried GP | 25% (n=3) | Locum and salaried GP | 33% (n=3) |
| Participant identifier | GP1-GP12 |  | *Focus group 1 (n=4)* | *Focus group 2 (n=5)* |
|  |  |  | GP1, GP9, GP10, GP14 | GP2, GP3, GP5, GP11, GP12 |

### Principal findings

We found that the key facilitators for GPs to change their behaviour from environmentally less sustainable to environmentally more sustainable during consultations were facilitators that motivated the provision of sustainable care through a focus on the benefits [*TDF: beliefs about consequences; COM-B: motivation*] and the opportunities created by the physical and social environment [*TDF: environmental context and resources* and *social influence; COM-B: opportunity*]. Additionally, for almost all participants, the starting point of the change did not lie within these domains, but was a moment of realisation, when the impact of healthcare on the planet sank in *[TDF: knowledge; COM-B: capability*]. Figure 1 gives a visual representation of these general findings.

**Figure 1.**
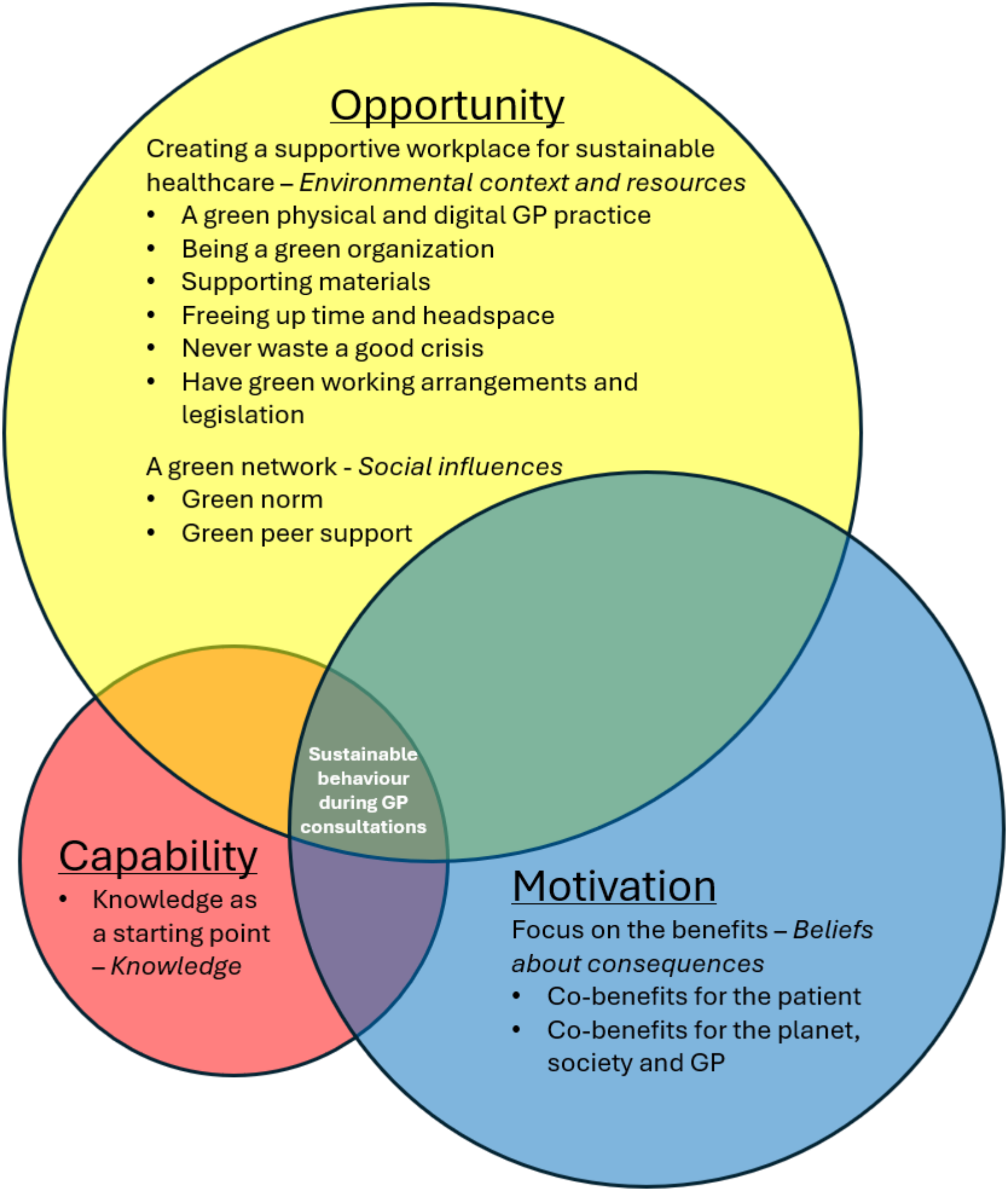
Key facilitators for green behaviour change for GPs during consultations, according to TDF (Theoretical Domains Framework; italics)(16) and COM-B model (underscored).(17)

An overview of all facilitators, organised per TDF- and COM-B domains, is provided in table S2, and supplement 6 presents detailed descriptions and illustrative quotes for each facilitator.

### Facilitators for sustainable behavioural change during consultations

#### Knowledge as a starting point

> *[COM-B: capability; TDF: Knowledge]*

Nearly all participants described a moment of realisation, after which they began to change their behaviour towards more sustainable healthcare. This was knowledge about the impact of healthcare on the planet, but also knowledge about sustainability, e.g. plant-based diets, in their personal lives that found its way into their professional life.

> *GP5: ‘And then I suddenly had a sort of ‘eureka’ moment where I realised, “Hey, I’m on that GP training programme and, actually, we’re not doing anything at all about sustainability*.*”’*

#### Focus on the benefits

> *[COM-B: motivation; TDF: Beliefs about consequences]*

### Co-benefits for the patient

The main benefit participants elaborated on, was the co-benefit for the personal health of their patients, both in the short and long term. They reported that they tried to make a connection to the personal health of their patient, and that that is the main argument to opt for the sustainable option. For example, when opting for lifestyle interventions instead of medication, they focus on the self-efficacy of the patient and the prevention of potential side effects of medication; or focusing on the importance of clean air with patients with lung disease.

> *GP11: “I always try to see it from their [the patient’s] perspective, you know? ‘What’s in it for them’ if we’re going to try this*.*”*

Furthermore, it also helps to focus on the practical co-benefits for the patient, such as easier treatment adherence and more tailored medication packaging which reduces the amount of medication at home.

Lastly participants stretched that when proposing a more sustainable alternative, effective communication with the patient, and as described above focussing on the co-benefits for the patient, is a key to success.

### Co-benefits for the planet, society and GP

The primary benefit mentioned by participants was the intended positive impact on the planet, including reduced CO_2_ emissions and less waste. Societal co-benefits included potential healthcare cost reductions through assumed cost-effective lifestyle interventions. Lastly, the participants emphasized the two co-benefits for them as GPs. First helping them to be able to practise their profession better because planetary boundaries helped them to deliver tailored healthcare. Second, they experience a reduction in workload as they need to prescribe fewer medicines or request fewer tests.

> *GP6: “Yes, one fewer diagnostic request simply means one less thing to do”*

### Creating a supportive workplace for sustainable healthcare

> *[COM-B: Opportunity; TDF: Environmental context and resources]*

Participants indicated that working in a supportive workplace, helped to change their behaviour; which may be split up into six subthemes.

### A green physical and digital GP practice

A green physical and digital GP practice encompasses references to sustainability both within the GP practice itself, e.g. infographics on sustainable healthcare, but also a statement on the website. These expressions serve both as a reminder to the GP and colleagues to provide sustainable healthcare, but also to legitimise the provision of sustainable healthcare to patients.

> *GP10: “Well, at my practice, we had a statement on our website explaining that we highly value sustainability—what that means to us and what we’re doing about it—and we also displayed a message on the waiting room screen. That way, patients know you’re actively working on it, and it also legitimizes the fact that you’re incorporating it into your consultations. I think that really helps too*.*”*

GP software is also viewed as supporting the delivery of sustainable healthcare, by making it easier to choose the sustainable option by defaulting the sustainable option or providing sustainability information (e.g. number of tablets per package for less spillage), or by making it insightful which patients use chronic medications from the repeated prescriptions list. In the future, it is also seen as helpful to have the carbon footprint of medication in the prescribing system.

### Being a green organization

It helps to have an organization culture that supports sustainable healthcare. For example, by fostering a sense of shared ownership of the sustainable mindset by assigning sustainability projects to all staff members. But also, by drawing on colleagues’ skills or expertise. For example, a practice nurse who has more time for, and experience with, discussing lifestyle interventions. Lastly, it helps to have recurring actions that help sustainable healthcare within a practice, e.g. by carrying out a regular medication review for selected patients to discontinue unnecessary medication.

### Supporting materials

#### Green guidelines

There are several ways that incorporation of sustainability into guidelines helps. For the green GPs in our sample the green guidelines serve as a norm setting document when discussing sustainable healthcare with colleagues.

> *GP11: “Well, what I particularly like is that you can also justify things to your colleagues, can’t you? You can just say, ‘Look, it’s in the guidelines too*.*’”*

Additionally, participants stated that they cannot know everything, it is therefore beneficial that sustainable healthcare is already incorporated in the guidelines. Lastly, it helps GPs and GP trainees to learn more about sustainable healthcare.

#### Patient-specific materials

Using patient-specific supporting materials helps to deliver more sustainable healthcare, as it helps to convince patients to opt for the sustainable option, for example using the Lifestyle Wheel (‘Leefstijlroer’).(22) Information on sustainable healthcare on patient medical information websites is also seen as helpful in the future. In addition, the use of devices such as the In-Check DIAL is also seen as helpful;(23) this can assist in determining whether someone is suitable for switching to a sustainable alternative.

> *GP12: “[The In-Check DIAL] that really did help our practice nurse a great deal. [*…*] To convince people that: ‘This [a DPI] is definitely just as good for you. […]’”*

### Freeing up time and headspace

Within GP consultations, but also within a GPs workweek it helps to free up time for sustainable healthcare. Within consultations this time helps to better explain the different options, sustainable healthcare, or to discuss lifestyle changes. During the workweek, deliberately allocating dedicated time to sustainable healthcare, for example half a day per week, can reduce the perceived burden of integrating sustainability into daily practice and support its long-term implementation. Thirdly, it helps to allocate time to learn more about sustainable healthcare, for example by planning a sustainable day each month to have a seminar about sustainable healthcare or have Green Pharmacology meetings.

### Never waste a good crisis

Making the most of unexpected events that temporarily necessitate the provision of sustainable healthcare helps to ensure that more sustainable healthcare is provided in the long term. For example, the shortage of pMDI’s during the winter of 2024 helped to switch more patients to the more sustainable alternative, the DPI.

### Have green working arrangements and legislation

It also helps to have green working arrangements with partners at the neighbourhood level, with the pharmacist as the key partner. For example, pharmacists may select an equivalent DPI based on stock availability, provided this makes no difference to the patient, to prevent medication expiry.

Lastly, participants feel supported when existing regulations align with sustainable healthcare practices. For example, currently Dutch dispensing and reimbursement policies already limit the medication quantity supplied upon first prescription to a 15-day supply.

#### A green network

> *[COM-B: Opportunity; TDF: Social influence]*

### Green norm

Providing sustainable healthcare is increasingly seen as the norm in general practice, that makes it easier to follow suit.

Locally in individual GP practices sustainable healthcare may be the norm through daily practice. These include for instance, opting for sustainable treatment options where possible, but also through business operations by opting primarily for second-hand items.

> *GP6: “If, for example, you look at how they [colleagues*.*] already sort their waste properly, then it is certainly easier to go along with that*.*”*

This green norm also applies to General Practice as a whole. Participants indicate that making green choices already felt as the norm for GPs, as the non-medication option, often the more sustainable option, has always been the first step in Dutch GP guidelines.

Furthermore, sustainable healthcare is also receiving increasing attention at a societal level, for example in the media. As a result, sustainable healthcare now seems to be increasingly becoming the social norm, making it easier to put into practice in the consulting room.

This is supported by the perception that patients also view a focus on non-medication advice or lifestyle changes as standard practice.

### Green peer support

Working together as a team within a GP practice and local community makes it easier to deliver sustainable healthcare. This lowers the bar to propose sustainable actions and evokes a sense of belonging.

Furthermore, it helps to have the social support of other green GPs. Within a local GP network, but also on a national level. This makes that you feel less as the odd one out, or a voice crying in the wilderness.

> *GP5: “I remember that back then, during those first few meetings [*…*] you really thought: ‘Wow, there are others out there working on this too*.*’”*

#### Helpfulness and feasibility

All actionable facilitators scored highly in both focus groups in terms of helpfulness and feasibility with an average of 3.6 [2.6-4.0] and 3.2 [2.3-3.9] respectively on a 1–4 Likert scale. The visual representation and related scores of the prioritised facilitators can be found in supplementary figure S1.

## Discussion

### Summary

In this study we identified key facilitators for GPs to change their behaviour from environmentally less sustainable to environmentally more sustainable during consultations. We found that the starting point for behaviour change was a moment of realisation, when the impact of healthcare on the planet sank in. The facilitators sustaining the change were ones that motivated the provision of sustainable care through a focus on the benefits, and the opportunities created by the physical and social environment.

### Strengths and limitations

By applying the behavioural sciences lens, combined with the expertise from general practice, sustainable healthcare and qualitative research, we have been able to zoom out from specific behaviours and unravel the pattern driving the behaviour change. By drawing on the experiences of pioneers, we have overcome the mindset of focusing on barriers and shifted our focus to success stories. This makes it possible to design interventions based on what actually works.

We also recognise limitations to our study. First, the focus groups were designed in line with a toolkit and planned to give prioritization to the found facilitators but proved to add less to the data than we aimed. Future studies should consider shifting to a more general population for prioritisation, to avoid a cluster of highly scored facilitators. Secondly, our focus on Dutch GPs may limit the transferability of our findings to other primary care contexts. To account for this, we used the TDF, which examines the underlying patterns of behaviour, to identify patterns that may prove relevant in these settings too. Lastly, our focus on green GPs, who are pioneers in this field, may have downplayed the potential side effects perceived by the wider GP population, such as concerns about increased workload. However, our findings suggest that these concerns are not necessarily inherent to sustainable GP care itself. Participants even reported a decrease in workload as they had to prescribe less and do less diagnostics.

### Comparison with existing literature

#### Principal findings – the underlying pattern

By zooming out from specific behaviours to the overarching domains of behavioural change, an underlying pattern emerges. This pattern of TDF domains largely aligns with findings from previous TDF-based reviews within sustainable healthcare.(24, 25) In these reviews, environmental context and resources, social influence and knowledge are also identified as key domains. Our findings provide more detailed and consultation-specific insight into these domains for GPs by illustrating how change can be initiated through moments of realisation and facilitated through opportunities in both the physical and digital work environment, as well as through social support and norm-setting among colleagues.

Additionally, our findings add an extra dimension by emphasizing that beliefs about consequences also play an important role for GPs during consultations, through focusing on the benefits of sustainable healthcare. These benefits of sustainable healthcare for patients are also endorsed by studies among Irish and Danish primary care professionals, including GPs,(5, 26) and Dutch cardiologists.(27) In addition to these healthcare professionals’ views, patients also consider the benefits to their own health to be important in the transition to sustainable healthcare.(28) The approach taken by the ‘green’ GPs is thus in line with patients’ wishes regarding sustainable healthcare, and may therefore even strengthen the patient-doctor relationship that healthcare professionals feared they might lose.(13, 14)

#### Knowledge as a starting point, not as the sole facilitator

Besides the previous mentioned concerns about the doctor-patient relationship, GPs have also identified a lack of knowledge as a barrier to implementing sustainable healthcare.(13, 29) While our findings support the importance of knowledge in triggering behaviour change, they suggest that knowledge alone is insufficient. Rather, sustained change appears to be driven by environments that facilitate sustainable behaviours and by the benefits that GPs experience.

#### Co-benefits of Sustainable Healthcare for Healthcare Delivery

Focusing on the benefits of sustainable healthcare is a valuable facilitator for behaviour change. These benefits may extend beyond the elements already mentioned, particularly when GPs use the co-benefit of delivering more appropriate care to tackle overmedicalisation. Overmedicalisation is viewed by GPs as a problem, and a barrier to sustainable healthcare.(5, 29) However, our participants indicated that considering planetary boundaries actually supports them in delivering more appropriate, high-value care. As such, framing sustainable healthcare as a means of delivering better care may help reduce overmedicalisation while simultaneously advancing sustainability. According to the 60-30-10 rule (60% high-value, 30% low-value, and 10% harmful care), this approach could potentially avoid up to 40% of low-value and harmful care, together with its corresponding carbon footprint, while maintaining or even improving patient health outcomes.(30, 31)

### Make it easy

The last two key facilitators include making good use of the physical, and social environment, i.e. making it easier to adopt more sustainable behaviour. This ties in with the findings of Irish and German studies in which primary care professionals were asked about sustainable healthcare.(6, 26) They indicated that they fall back on familiar processes and workflows when the workload becomes too high, and that easing sustainable healthcare into these processes helps. How the environment can be utilised to promote sustainability is showcased by Canadian GPs, who phased out exam table paper in their practice. Their findings show that not only the use of exam table paper decreased, but – in line with our results – the staff’s environmentally friendly behaviour increased.(32)

Furthermore, our findings and those of Walsh et al.(26) suggest that providing carbon footprint information within GP software could support more sustainable prescribing decisions. This requires reliable and comparable environmental impact data. To this end, scientists have called on the EU to adopt a standardised method for calculating the environmental footprint of medicines.(33) The value of such information is illustrated by a CE Delft report comparing methods for cleaning medical instruments in general practice,which translated environmental impact data into practical recommendations that GPs can implement with minimal effort.(34)

#### Implications for research and practice

As we now know the domains of behaviour change that encompass the key facilitators, we can draw on specific behaviour change techniques to design and study appropriate interventions.(35) This process is already used for the sustainable switch from liquid antibiotics to solid antibiotics, which proved successful.(36, 37) Future research can focus on this through general changes to the GPs’ work environment. This could include altering the choice architecture used in GPs prescription software, for example, by adding environmental labels to medication or other nudges, such as using the default option, in cases of medical equipoise.

Furthermore, our findings can help to teach GPs and GP trainees, but also medical students to adopt more sustainable behaviour. For example, an introductory lecture or webinar on planetary health and sustainable care could serve as a moment of realisation. The focus herein should not only be the urgency of the ecological crises but should also emphasize on the benefits of sustainable healthcare. Yet, it is important that the environment in which GPs or students subsequently practise is conducive to the provision of sustainable care. Setting an assignment aimed at improving this environment, such as a Sustainability Quality Improvement (SusQI) project,(38) may be helpful in this regard.

## Conclusion

Changing GPs’ behaviour from environmentally less sustainable to environmentally more sustainable during consultations begins with realising the impact of healthcare on the planet, and is sustained by focusing on the benefits of sustainable healthcare and creating opportunities within physical and social environments. By building on these facilitators, future research and practice should focus on altering care environments to enable GPs to easily opt for the sustainable option.

## Data Availability

The protocol and logbook will be available on the Open Science Framework after publication. Raw data is not available to ensure privacy of our participants.

https://osf.io/z76jf

## Ethical approval

The study was approved by the non-WMO committee of the Leiden University Medical Centre (#25-3036).

## Data

The study protocol and logbook will be available on the Open Science Framework after publication (https://osf.io/z76jf).

## Acknowledgments

The authors would like to thank the participating GPs for contributing their time and perspectives, and dr R. van der Kleij, for her contribution to the design of the study.

## Use of Artificial Intelligence

During manuscript preparation, the authors used Microsoft Copilot to assist with language editing and text refinement. All suggestions were critically reviewed, verified, and, where appropriate, revised by the authors. The authors take full responsibility for the final content of the manuscript.

