## Supplementary data for "Facilitators for green behaviour change during GP consultations: a qualitative study"

#### 1. Table S1

| No and item | Description |
|---|---|
| **Domain 1: Research team and reflexivity** | |
| *Personal Characteristics* | |
| 1. Interviewer/facilitator | Interviews: EV; Focus groups: IS (moderator) and EV (observer) |
| 2. Credentials | EV: MD; IS: PhD; SK: MSc |
| 3. Occupation | EV: PhD candidate and GP in training; IS: senior researcher; SK: PhD candidate |
| 4. Gender | EV: female; IS: female; SK: female |
| 5. Experience and training | EV: trained as a medical doctor, and during PhD trajectory in qualitative research, this was her first full qualitative study; IS: trained as a nurse and education specialist, and had training in and extensive experience with qualitative research; SK: trained as a psychologist, and currently undertaking a PhD in behavioural sciences |
| *Relationship with participants* | |
| 6. Relationship established | Prior to her research, EV had already been involved in sustainable healthcare as a volunteer and knew two of the participants through this. During her research into sustainable GP care, EV gave regular presentations and had already met two of the participants on these occasions. One participant already knew EV from her medical studies. EV did not know the other participants. |
| 7. Participant knowledge of the interviewer | The participants may have been aware of EV's (previous) work on sustainable healthcare through earlier publications or media coverage. From this, they might have concluded that EV considers making healthcare more sustainable to be important. |
| 8. Interviewer characteristics | The involvement in sustainable healthcare and familiarity with the network mentioned above, and the potential impact of these factors on the data and results, are described in the methodology section of the article. |
| **Domain 2: study design** | |
| *Theoretical framework* | |
| 9. Methodological orientation and Theory | Interviews: A realist approach was adopted. Data were analysed using a hybrid deductive-inductive thematic analysis, combining TDF-based coding with inductive theme development within domains. (1, 2) |

| | |
|---|---|
| | Focus groups: Focus groups were conducted using the Prioritizing Implementation Barriers toolkit and analysed using descriptive statistics.(3) |
| *Participant selection* | |
| 10. Sampling | We recruited participants through purposive sampling (variation: gender, age, experience as a GP and with sustainable healthcare, and workplace type), and snowballing. |
| 11. Method of approach | We approached participants using a messaging platform (Signal Messenger LLC, Mountain View, CA, USA) or email. |
| 12. Sample size | We approached 32 green GPs and included 13 green GPs (interviews n=12; focus groups N=8 from interviews, n=1 new participant). |
| 13. Non-participation | 19 participants approached through membership of 'Groene Huisarts'(n=10), snowball sampling (n=6), or self-selection (n=3), declined participation. They either did not respond, or reported being unavailable during the data collection period, too busy, or unable to attend the proposed appointment times. |
| *Setting* | |
| 14. Setting of data collection | Participants were given the option of online interviews or face-to-face interviews at a location of their preference. All participants opted for an online interview via Microsoft Teams. Both focus groups were conducted via Microsoft Teams. |
| 15. Presence of non-participants | N.a. |
| 16. Description of sample | The majority of the participants were female (around 80%). The years of experience in general practice was mainly double the years of experience with sustainable healthcare. All types of workplaces for a GP were represented. |
| *Data collection* | |
| 17. Interview guide | Interviews: an interview guide was developed based on the vignettes and questions were designed in line with the Theoretical Domains Framework to subtly prompt participants if needed. The interviews were semi-structured. The content of the vignettes was based on a small pilot study using a questionnaire among a group (n > 260) of green oriented Dutch GPs and were written using Atkins et al.'s specificity rules. (1)<br>Focus groups: the topic guide was developed based on the Prioritizing Implementation Barriers toolkit.(3) |
| 18. Repeat interviews | No interviews were repeated |
| 19. Audio/visual recording | The interviews and focus groups were audio-recorded using Microsoft teams and an external audio recorder |
| 20. Field notes | Yes, field notes were taken during and after data collection. |

| | |
|---|---|
| 21. Duration | The interviews lasted on average 49 minutes [range 35 – 58]. The focus groups lasted 56 and 60 minutes, respectively. |
| 22. Data saturation | Data saturation was discussed during the design of the study and defined and estimated as follows in the study protocol: For the qualitative interviews we expect to have 10-15 interviews. We will continue interviewing until data-saturation is achieved.(4) In this study we define data saturation as the point at which no new topics or information emerge from an interview. When this point is reached, we will conduct one additional interview to confirm data saturation.<br>Data saturation was achieved after 12 interviews in total. |
| 23. Transcripts returned | No, transcripts were not returned to participants |
| **Domain 3: analysis and findings** | |
| *Data analysis* | |
| 24. Number of data coders | Two coders (EV and SK) and a third reviewer available (MA) |
| 25. Description of the coding tree | An a priori coding framework based on the 14 domains of the Theoretical Domains Framework (TDF) and their domain descriptions as reported by Atkins et al. was used for deductive coding. |
| 26. Derivation of themes | Following deductive coding to the TDF domains, themes were developed inductively within each domain by grouping related codes into theme piles, consistent with Braun and Clarke's approach to thematic analysis.(2) |
| 27. Software | We used Atlas.ti 9 (ATLAS.ti Scientific Software Development GmbH, Berlin, Germany) |
| 28. Participant checking | No, participant did not provide feedback on the findings. |
| *Reporting* | |
| 29. Quotations presented | Yes, participant quotations are presented to illustrate the facilitators, accompanied by a participant identifier. |
| 30. Data and findings consistent | Yes. Themes were supported by illustrative participant quotations and were grounded in the interview data. |
| 31. Clarity of major themes | Four overarching themes describing facilitators of sustainable GP care were identified and presented. |
| 32. Clarity of minor themes | Minor themes were omitted due to word count constraints but are available in a supplementary file. |

*Table S1 – Consolidated criteria for reporting qualitative research (COREQ) table(5)*

#### 2. Vignette design – pilot study

To design the vignettes, we conducted a small pilot study to identify green target behaviours. We aimed for a sample of Green GPs. We recruited participants using special interest groups on sustainable GP care on a messaging platform (Signal, Messenger LLC, Mountain View, CA, USA).

We carried out the pilot study using Castor software (Castor EDC B.V., Amsterdam, the Netherlands). We asked the participants to give a maximum of ten examples of behaviour changes from environmentally less sustainable behaviour to environmentally more sustainable behaviour during consultations. After the participants gave these examples, we asked them to rate the behaviour changes from:

1. Least to most applied new ecologically more sustainable behaviour
2. Least to most impactful new ecologically more sustainable behaviour in their opinion

From these answers we selected the three most applied and/or most impactful behaviour changes to discuss during the qualitative interviews. We wrote the vignettes using Atkins et al.'s specificity rules. (1)

At the end of the questionnaire an open text box was available, in which a participant could indicate whether they would like to be contacted for a follow-up qualitative interview and focus group and through which details (e-mail address or phone number) they could be reached.

### 3. Interview guide

**Introduction**

- **Informed consent on audio**
- **Brief introduction by interviewer EV:** name, affiliation
- **Goal:** The aim of the study is to gain an understanding of how GPs have changed their behaviour during consultations from less sustainable to more sustainable practices, and which factors have contributed to this.
- **Duration**: The interview is likely to last 30–60 minutes
- **Confidentiality and audio recorder**
    - *If conducted in person*: The interview will be recorded using two audio recorders and transcribed in full. Where possible, the audio files and transcribed interviews will be stored anonymously; otherwise, they will be stored under a pseudonym (e.g. GP1, GP2, etc.) on a secure drive at the LUMC. We intend to use the data obtained for a scientific publication.
    - *If conducted online*: The interview will be recorded via Microsoft Teams and an additional audio recorder and will be transcribed in full. Where possible, the audio files and transcribed interviews will be stored anonymously on a secure LUMC drive; otherwise, they will be stored under a pseudonym (e.g. GP1, GP2, etc.). We intend to use the data obtained for a scientific publication.
- **There are no right or wrong answers**
- **Did you sign the digital informed consent form?**
- **Questions?**

| |
|---|
| **General questions**<br>- How old are you?<br>- What is your gender?<br>- How many years have you been working as a GP?<br>- In what capacity do you work as a GP? (Locum, salaried GP, GP partner)<br>- How many years have you been actively engaged in sustainable healthcare? |
| We are going to look at three examples of sustainable behavioural change during consultations. These are specific examples, as we have found that it helps to focus on a particular example.<br>Using these examples, we will discuss which factors contributed to the behavioural change.<br>*Show three vignettes* |
| Vignette 1 – Sustainable prescribing of inhalation medication<br>As a GP, I used to prescribe a pressurised metered-dose inhaler for all lung patients with shortness of breath. Now I prefer to prescribe a dry powder inhaler. I only prescribe a pressurised metered-dose inhaler where there is a clear indication, such as for young children or patients with reduced inhalation strength.<br><br>Vignette 2 – prescribing less per consultation |

As a GP, I used to prescribe antidepressants for several weeks to patients who presented with persistent symptoms of depression. Now, when I start prescribing, I prescribe a smaller number of antidepressant tablets.

Vignette 3 – Focusing on lifestyle changes before starting preventive cardiovascular medication

As a GP, I used to automatically prescribe cholesterol-lowering medication for patients with an increased risk of cardiovascular disease. Now I first offer lifestyle advice, such as eating a healthy diet, taking more exercise and giving up smoking.

- Which example do you identify with the most?
- What factors helped you to change this behaviour?

*After this, there will be an open, exploratory discussion about these factors.*

Which other example do you identify with?

Are there any other factors relating to this behavioural change that helped, which we haven't discussed yet?

The domains of the **TDF** with optional prompting questions (only to subtly prompt the participant if needed):

*Knowledge*

- To what extent did knowledge about sustainable care contribute to a change in your behaviour?
  - Was there any specific knowledge that clearly contributed to this?
  - Where did you acquire this knowledge?

*Skills*

- To what extent have some of the skills you possess contributed to the change in your behaviour?
  - Which skills were most valuable to you in this regard?
  - Where did you learn these skills?

*Social/professional role and identity*

- To what extent do you consider promoting more sustainable behaviour in the consultation room to be part of your role as a GP?
  - Why do you regard this as part of your role as a GP?
- To what extent do you feel it is your responsibility to be more sustainable during consultations?

*Beliefs about capabilities*

- To what extent were you confident that you would be able to change your behaviour to be more sustainable?
  - How easy do you find it to adopt this more sustainable behaviour?

*Optimism*

- To what extent is it feasible to put this more sustainable behaviour into practice during consultations?

Beliefs about consequences

- What (positive) consequences does this more sustainable behaviour have for you?

| |
|---|
| ◦ To what extent do you think this more sustainable behaviour makes a positive contribution to the environment and the planet?<br>◦ To what extent do you think this more sustainable behaviour affects your doctor-patient relationship? In what way?<br>Reinforcement<br>- To what extent do you benefit from adopting more sustainable behaviour?<br>◦ If you do benefit, what is that benefit?<br>Intentions<br>- To what extent were you/are you motivated to adopt this more sustainable behaviour?<br>◦ Do you think you will still be applying this behaviour in 10 years' time?<br>Goals<br>- To what extent is adopting more sustainable behaviour in the consultation room important to you?<br>Memory, attention and decision processes<br>- To what extent does being able to easily recall this sustainable behaviour help you to actually put it into practice?<br>◦ Do you remember to practise this sustainable behaviour during every consultation?<br>◦ Are there any cues that help you remember to practise it?<br>Environmental context and resources<br>- To what extent do aspects of your consultation room or practice help to make it easier to adopt sustainable behaviour?<br>◦ What contributes the most?<br>Social influences<br>- To what extent does your patient's opinion influence the adoption of more sustainable behaviour?<br>- To what extent does your colleagues' opinion influence the adoption of more sustainable behaviour?<br>◦ Do you think your colleagues also adopt this more sustainable behaviour in practice?<br>Emotion<br>- To what extent do the emotions you experience when adopting more sustainable behaviour influence your actual adoption of that behaviour?<br>◦ Which emotions have contributed most to this?<br>Behavioural regulation<br>- Would it help if this more sustainable behaviour were part of the GP guideline?<br>- Do you have a way of monitoring how often you practise this more sustainable behaviour? If so, what is it? |
| **Closing remarks** |
| Are there any other points that haven't been discussed yet? Thank you for your participation. |

### 4. Topic guide focus groups

**Introduction**

- **Informed consent** review
- **Did you sign the digital informed consent form?**

**START RECORDING**

- **Focus group contents**
- **Introduction**
  - **Study aim**
  - **There are no right or wrong answers**
  - **Introductions (Moderator and observer; Participants)**
- **A glance back at the interviews**
- **Focus group explanation**
  - **Aim**
  - **Method (WooClap and visual presentation)**
- **Questions?**

**Prioritizing facilitators**

Using a digital tool, we would like to ask you to prioritise the identified factors that contribute to behavioural change from less sustainable to more sustainable practices during consultations, based on how helpful the factor was or is to you and how feasible it was or is. We would then like to ask you to discuss why each factor was helpful or feasible for you, in order to identify the most helpful and most feasible factors.

[Display link or QR code for the digital tool via WooClap and Excel]

**Displaying a 2x2 table showing the prioritisation of facilitators**

[Example of a 2x2 table from the Prioritizing Implementation Barriers toolkit]

Figure 1. Graph of importance and feasibility ratings.
"• = scored ratings"

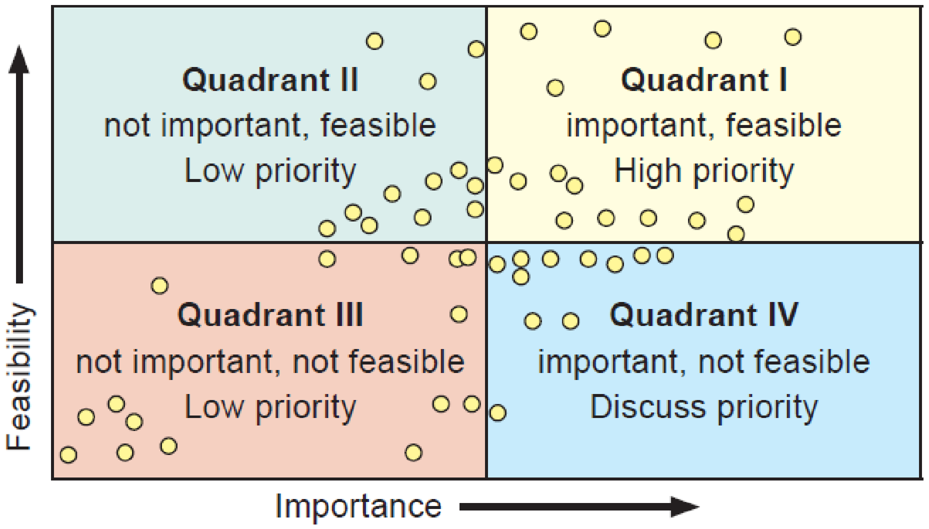

](3)

1. To what extent do you, as a GP, find it **helpful** in applying more sustainable behaviour during consultations to: [facilitator, e.g. approach each patient as a new opportunity to raise the issue of sustainability]?

| |
|---|
| a. *Helpful (1 = not helpful; 2 = somewhat helpful; 3 = helpful; 4 = very helpful)*<br>2. To what extent do you, as a GP, find it **feasible** to apply more sustainable behaviour in the consultation room by: [facilitator, e.g. approaching each patient as a new opportunity to raise the issue of sustainability]?<br>a. *Feasible (1 = not feasible; 2 = somewhat feasible; 3 = feasible; 4 = very feasible)* |
| **Discussion** |
| The table shows that factor X, factor Y, factor Z [...] are the most helpful and feasible for you.<br>Possible guiding questions for the moderator:<br>- Are these the right factors for GPs who are starting to change their behaviour during consultations to start with?<br>- Are there any differences of opinion amongst you as participants?<br>- Are there any factors elsewhere in the table or graph that require discussion?<br>- Which of the prioritised factors should be given the highest priority, and why?<br>- How can GPs who are starting to make behavioural changes towards more sustainable practices best apply these factors?<br>[Feedback from the moderator: So, I understand that, given the importance of these factors and their feasibility, you would opt for factors X, Y, Z, A and B for your fellow GPs. Is that correct?] |
| **Closing remarks** |
| Are there any other points that haven't been discussed yet? Thank you for your participation. |

#### 5. Table S2

| COM-B | TDF domain | Facilitators<br>*To deliver more sustainable healthcare it helps to* |
|---|---|---|
| *Capability* | Knowledge | Have knowledge about sustainable healthcare |
| | | Have awareness of existence (a realisation moment) * |
| | | Know where to get information from |
| | | Teach about sustainability |
| | | Use a green lens during consultations † |
| | Skills | Use guidelines and facts as a base for communication with colleagues |
| | | Have good communication skills with patients |
| | | Have experience as a GP |
| | Memory, attention, and decision processes † | Keep it manageable |
| | | Keep sustainable healthcare top-of-mind |
| | Behaviour regulation | Use self-monitoring |
| | | Prepare for sustainability conversations |
| | | Pick your battles wisely |
| | | Make sustainable healthcare a habit |
| | | Make it a project |
| *Opportunity* | Environmental context and resources § | Have a Green GP practice |
| | | Use the possibilities of GP software |
| | | Use patient information materials |
| | | Use supporting devices |
| | | Have a supportive organisational culture |
| | | Leverage the strengths and opportunities of colleagues |
| | | Have sustainability in guidelines |
| | | Have supportive legislation |
| | | Make use of unexpected events |
| | | Time |
| | | Use green working agreements |

| | | |
|---|---|---|
| | Social influences | Have sustainable healthcare as the norm within our GP practice |
| | | Have sustainable healthcare as the norm for GPs ‡ |
| | | Have sustainable healthcare as the norm for my patients |
| | | Have sustainable healthcare as the norm in society |
| | | Have social support in our GP practice § |
| | | Have social support from network partners |
| | | Have social support from like-minded GPs |
| *Motivation* | Social/-professional role and identity ‡ | See sustainable healthcare as part of a GPs job |
| | | Identify as a Green GP |
| | | Identify as a Green person |
| | Beliefs about capabilities ‡ | Make a connection with your patient |
| | | Have good communication skills |
| | | Start with small sustainable actions and build from there |
| | | Believe in sustainable healthcare |
| | Optimism | Optimism |
| | Beliefs about consequences ‡ | Focus on the benefit for the planet |
| | | Focus on the co-benefit for the patient |
| | | Focus on the co-benefit for society |
| | | Focus on the co-benefit for me as a GP |
| | Reinforcement | Make being more sustainable a game |
| | | Focus on successes |
| | | Get positive feedback from patients |
| | | Reframe negative experiences into motivation to try harder ◊ |
| | Intentions * † | Feel committed to sustainable healthcare |
| | Goals | Set goals |
| | Emotions ◊ | Enjoy delivering sustainable healthcare |

*Table S2 – Facilitators for green behaviour change in clinical consultations, grouped by the COM-B model and per corresponding TDF domain; with cross-references to related domains.*

### 6. Explanation of all facilitators with related quotes

#### 1. *Knowledge*

##### A. Have knowledge about sustainable healthcare

Almost all participants cite knowledge as the starting facilitator for their behaviour change towards greener practices in the consulting room.
*GP10 (10:28) "[...] it's all about knowledge, including, for example, understanding how the guidelines are structured. [...] And, of course, having the knowledge about how this affects the climate and the planet."*
Participants shared specific information that helped them, or they think will help them to do even more sustainable in the future. They talked about practical knowledge which may be found in their working environment such as knowing the quantity of a medication package and knowing the CO2 emissions of medication or doing diagnostics. But also, the knowledge of how to translate this abstract $CO_2$ emission figure into a more understandable format, such as kilometres travelled by car or airplane. Lastly, general GP knowledge such as knowledge about different types of inhalation medication and their pro's and cons, and knowing of alternatives for climate unfriendly medication options.
*GP1 (1:68): "How much CO2 does that [X-ray, red] actually produce?"*

##### B. Have awareness of existence (a realisation moment) *

Participants shared that there was a point at which they first received information about sustainable healthcare, which elicited two types of response. Some participants indicated that from the moment they first learnt about the impact of healthcare on the planet, they immediately made significant changes to their behaviour to provide more sustainable care. Others indicated that for them it is more of a gradual process in which they are delving deeper and deeper into sustainable care; this arouses their curiosity and inspires them to explore the subject further.
In addition, there is also a group of participants for whom their interest in sustainable healthcare originated from a personal interest in their personal lives, such as having knowledge about plant-based diets.
*GP5 (5:6): "[...] then you learn that, well, those things aren't actually that good, and so you start to look at things much more critically and ask, 'OK, which patients actually need this?"*

##### C. Know where to get information from

Participants shared various ways that they gathered the knowledge that helped to be more sustainable. This may be divided into three categories: individual (i.e. podcasts, articles, guidelines), in groups (i.e. continuing professional development (CPD) courses, talks on sustainability, webinar, lecture, pharmacological meeting on sustainability) and during formal education within GP specialty training.

##### D. Teaching about sustainability

Explaining about sustainable healthcare to others through lectures or other teachings, helps to better understand sustainable healthcare yourself.

*GP6 (6:2) “But if you actually have to teach it, so to speak, then, well, then you really do get it [...]”*

##### E. Use a green lens during consultations †

Using the green perspective helps to be more sustainable during clinical consultations. This is the awareness of the products used and what’s actually needed. Additionally, it is the awareness of the origin of the raw materials until the full end product.
*GP7 (7:42) “That I really think about the raw material [...] and then, OK, that’s what a product is made from. “*

#### *2. Skills*

##### A. Use guidelines and facts as a base for communication with colleagues

When communicating with colleagues, many participants mention using the guideline as a basis for their arguments when discussing the choice of a particular treatment with a colleague. One participant notes that this allows them to justify why they have made a particular choice. Another participant also uses the guidelines as a tool to address a colleagues’ unsustainable choice, as well as in conversations about sustainable care with GP trainees.
It helps to use something tangible as a comparison: Using specific facts to underpin the message: for example, linking a medication switch to the reduction in kilometres driven in a gasoline car or a flight to a big city.
*GP9 (9:10) “Then you can simply say: ‘You know, the guidelines have changed in the meantime. Keep that in mind and please work in accordance with the guidelines.”*

##### B. Have good communication skills with patients

Good communication skills with patients help to deliver sustainable care during consultations. This involves various elements; if sustainability is not explicitly mentioned but the sustainable option is chosen, it helps to convey a sense of experience as a GP. Several participants also indicate that providing a clear explanation of the treatment is important for its success. Standard phrases that do not need to be specific to sustainable care can be used for this purpose. If sustainability is explicitly mentioned, conveying a sense of experience is also important to the participants. In addition, they use various communication strategies, such as gauging whether someone is open to information about sustainability or pointing out that we no longer do things that way (climate-unfriendly [ed.]) these days.
*GP 4 (4:10) “Look, with people who have never used inhalation medication before, you just say: well, I’m going to prescribe something that we know opens up the airways.”*

##### C. Have experience as a GP

Having experience as a GP, and having experience with sustainable care, builds self-confidence and thereby helps to put sustainable healthcare into practice. Participants say that it becomes easier once you have practised sustainable care a few times; this gives you confidence in your own abilities.
*GP4 (4:5) “[...] and then, once you’ve done it a few times, you’ll get the hang of it and it’ll go well.”*

##### 3. *Social and professional role and identity*

###### A. Set sustainable healthcare as the new norm ‡

If there is a change in the way you practise as a GP, e.g. if you become a GP partner, it is possible to establish a 'new' sustainable norm from the outset. This makes it easier to provide sustainable healthcare in the future.
*GP7 (7:26): [...] when I took over this practice, annual prescriptions were being issued. Our position is: no, we don't do that.*

###### B. See sustainable healthcare as part of a GPs job

Because providing sustainable care is seen as part of the GP's role, it is easier to put sustainability into practice. Sustainability is regarded as part of the profession because GPs collectively strive to promote health, and a healthy planet is a key prerequisite for this. For the participants, this involves not only prescribing in a more sustainable way, but also, for example, providing preventive care that helps to reduce health problems and healthcare use in the future.
*GP10 (10:23): but [sustainability] is actually just part of providing good GP care.*

###### C. Identify as a Green GP

Identifying as a 'green doctor' supports the delivery of sustainable healthcare.

###### D. Identify as a Green person

Several participants indicated that, at first, they were increasingly interested in sustainability in their private lives, and that this gradually extended to providing sustainable GP care. In their personal lives, this ranged from eating a plant-based diet at home to reducing waste.
Although the group of participants was selected as pioneers in the field of sustainable GP care, several actively expressed that they didn't identify as activists.
*GP2 2:4: [...] I've always lived as sustainably as possible at home, all my life. I'm just an eco.*
*GP9 9:62: Yes, because I certainly wouldn't describe myself as a huge activist – definitely not [...] I'm definitely trying my best. But I'm certainly no saint. And yes, I do try to get that message across: that you just, you know, as long as you do your best, every little thing helps.*

##### 4. *Beliefs about capabilities*

###### A. Make a connection with your patient

In various contexts, sustainable care can, alongside its benefits, also cause discomfort or a sense of loss for the patient. Participants indicate that by actively engaging in dialogue with patients on this subject and building a connection with them, it is possible to alleviate this discomfort or to take even a small step towards sustainability together with the patient.
*GP11 11:11: [...] Then I think, 'Right, so this is apparently something he doesn't agree with, or something.' Fine, that's that then, let's see where we can actually find common ground.*

##### B. Have good communication skills

When proposing a more sustainable alternative, communication with the patient about this is a key to success for many participants. This encompasses giving explanation about why you propose, what you propose; having confidence in conveying the message to the patient; managing expectations about the treatment and potential side effects.
*GP 5 5:12: I just use those powder inhalers, along with clear instructions, a proper explanation and good management of expectations.*
*GP4 4:28: [...] if I just repeat enough that it's really going to work and that I'm confident it will, then it'll all work out.*

###### C. Start with small sustainable actions and build from there

By starting with easy changes towards sustainable healthcare, participants indicated building confidence in their practice and stepping up towards more types of sustainable healthcare. This gave the feeling 'I can do this' or 'this is really easy', which helped lower the bar to be more sustainable throughout the working day. Which also translates into easier communication with patients, because you have more confidence in conveying the message.
*GP1 1:14; Because it's really easy, isn't it? So, whether you're prescribing one or the other, well, you're prescribing something anyway, so you might as well prescribe the right one. And yes, it takes relatively little effort.*

###### D. Set sustainable healthcare as the new norm ‡

Setting a new norm helps to convey the sustainable treatment option to the patient.
*GP5 5:15: In a way that really conveys the message: This is normal; it's standard practice. It's not some sort of extra difficult step. We're not going to do anything crazy just for the sake of sustainability. It's just normal.'*

##### E. Believe in sustainable healthcare

It helps to genuinely believe that you're doing the right thing; that makes providing sustainable care easier.

#### *5. Optimism*

Participants are optimistic that sustainable care will grow in the coming years.
*GP12 12:28: I do get the feeling that this will still be the case in time. In fact, I think that, in time, far more people will become more conscious of this – the conscious process of prescribing medication.*

#### *6. Beliefs about consequences*

###### A. Focus on the benefit for the planet

Participants stretch that it helps to know and feel that by delivering sustainable healthcare, there is less $CO_2$ emissions, less spillage, etcetera. Which then helps to lower the impact on our planet.
*GP4 4:9; I'm making this change because I think it's important – let's say – for the environment, and therefore, ultimately, for the future of humanity.*

##### B. Focus on the co-benefit for the patient

###### a. For their health

The main consequence participants elaborated on, was the co-benefit for the personal health of their patients, both in the short and long term. They said that they try to make a connection to the personal health of their patient, and that that is the main argument to opt for the sustainable option. For example, when opting for lifestyle interventions instead of medication, they focus on the self-efficacy of the patient and that in this way you can prevent the potential side effects medication can have. Or focusing on the importance of clean air with patients with lung disease.

*GP11 11:20: Yeah, I almost never bring up sustainability as an argument with patients, unless I know that's something they care about—otherwise, they'll just think, "Yeah, that's just you, isn't it?" Then it doesn't feel like something that matters to them anymore. I always try to see it from their perspective, you know? "What's in it for them" if we're going to try this.*

*GP9 9:16: And it's also related to the patients' symptoms, you see. These are patients who have trouble breathing, so they understand that clean air is important. Yes, so you can certainly make that connection as well. So those are all things that help.*

###### b. Practical

Participants describe that it helps to convey the patient to opt for a sustainable treatment option, by highlighting the co benefits it has for them. For example: 1) Participants stretch that when switching from a pressured metered dose inhaler (pMDI) to a dry powder inhaler (DPI), it also has the benefit for the patient that it's easier to use, easier to take with you, that it counts the number of inhalations, which all lead to a better adherence to the treatment. 2) When choosing a lifestyle intervention rather than medication as a treatment, it gives the patient more ownership over their health. 3) When prescribing is more tailored to what a patient already has at home, it's better for the environment but also leads to less unused medication boxes for the patient.

*GP8 8:37: [...] that I often find a disc [DPI] easier in terms of adherence to treatment, and that it's easy to take with you. And, and that it's also just less prone to mistakes in terms of how you use it.*

##### C. Focus on the co-benefit for society

Sustainable healthcare is seen as cost effective, which supports some participants in opting for the sustainable alternative.

##### D. Focus on the co-benefit for me as a GP

###### a. Less workload

Sustainable healthcare sometimes also means providing more tailored care or less care. This reduces a doctor's workload because there are fewer actions to do.

*GP6 6:44: Yes, one fewer diagnostic request simply means one less thing to do*

###### b. It helps me to be a good GP

Providing sustainable healthcare challenges GPs to consider their treatment options even more carefully. One participant expressed that it just helps to be a better GP. Others also told that trying to be sustainable supports them in not doing excess diagnostics or prescriptions.
*GP10 10:43: Take NSAIDs, for example – you really try to prescribe them as little as possible because they're not sustainable, but they're also simply not good for the patient. Most patients – certainly those with chronic pain – simply don't benefit from them, and that makes it easier to realise that prescribing fewer pills is actually almost always better for a patient. [...] You can also look at it the other way round: prioritising sustainability helps you be a good GP, so doing the right things, as you really should, don't you think? So it helps you to have that conversation in the first place, or to delay writing a prescription, or to say no. Even though the patient really wants some oxycodone or whatever, so it actually makes me better at practising my profession properly.*

##### E. Feel like you can change the norm ‡

By delivering sustainable healthcare one participant said that he aims to change the norm to be more sustainable. Others hope that by setting an example or giving tips via a newsletter they can inspire others to follow along.

#### *7. Reinforcement*

##### A. Make being more sustainable a game

If you track your progress in deprescribing for example or switching to a more sustainable alternative, you can make it a little game for yourself to improve the next time you monitor your GP practice.

##### B. Focus on successes

It makes the GPs have a success moment when the effectively switch patients to a more sustainable alternative or opt for the more sustainable alternative from the start. For example, when blood pressure is better regulated with a lifestyle intervention, this reinforces them to deliver sustainable healthcare in the future.
*GP1 1:56: Oh, those puffers do give me a sense of satisfaction, because they have an effect, don't they? So, because I'm the one who decides we're going to switch over; and because of my behaviour, you can see that it makes a big difference to the outcome.*
*GP2 2:58 Yes, when we'd got down to minus 100 at one point, we had a little party. [...] When we'd got down to minus 200 tablets, we had another little celebration. So, in the end, over the course of 1.5 years, I'd managed to reduce the number of pills taken by 270. Not just me, but my team as well, right? To be precise, and that's 365 days, so that's nearly 100,000 fewer tablets. [...] Yes, so if you do the maths, every pill counts, for 365 days, doesn't it? So, when you start tapering off chronic medication, you get a huge sense of satisfaction from it, because it produces really significant results very quickly.*

##### C. Get positive feedback from patients

It helps to get positive feedback from patients; this reinforces GPs to deliver sustainable care.
*GP10 10:40: No, I think it's just that it's really nerve-wracking to actually say it out loud, so you have to get past that hurdle, and if you then get positive reactions, that helps.*

##### D. Reframe negative experiences into motivation to try harder

It isn't always possible to opt for the sustainable option, which can make you feel as though you've 'let yourself down'. For some participants, this feeling actually encourages them to try again next time and choose the sustainable option.

#### 8. *Intentions*

##### A. Have a moment of realization *

Many participants describe some form of realization that the care they were giving to their patients wasn't sustainable. This led to curiosity in sustainable healthcare and evolved into becoming a Green GP.
*GP5 5:1 And then I suddenly had a sort of 'eureka' moment where I realised, 'Hey, I'm on that GP training programme and, actually, we're not doing anything at all about sustainability.'*

##### B. Be aware of sustainability †

Being aware of the impact a treatment has on the planet, helps to choose for a more sustainable alternative.
*GP4 4:29 Yes, it made me aware that, well, if you start something or commit to it for a longer period, you should also mention that they can eventually stop again once their body is back in balance or something like that. Something along those lines. So, I also mention when they can stop – I think that's something I'm now doing more actively.*

##### C. Feel committed to sustainable healthcare

It helps if you feel committed to sustainable healthcare.
*GP6 6:18 But then again, overall, I do want to commit to that, though more because I personally think it's important than because I feel it fits my role.*

#### 9. *Goals*

##### A. Set goals

Having clear that you want to be involved in sustainable healthcare, helps to behave accordingly.

#### 10. *Memory, attention and decision processes*

##### A. Deliberately consider the feasibility of sustainable healthcare †

It helps to be aware and actively question yourself whether it is possible to opt for the more sustainable treatment option for this patient.

**B. Keep it manageable**

GPs try to prevent cognitive overload, by deliberately choosing what to focus on and what not (e.g. focusing on CO2 emissions, rather than waste), by defining groups for which the sustainable option may be considered, by choosing a set moment during the workweek to actively work on sustainability with your patients and let it go for the rest of the week

*GP6 6:46 Waste is quite a big problem. But then again, you could solve your waste problem entirely, but with global warming of 3.5 degrees, you'd actually still be facing a really huge problem. So, in my view, emissions are actually a bigger problem. So, when I think about, say, a metered-dose inhaler – that's a real source of emissions – I feel that, personally, doing something about that might be more useful.*

**C. Keep sustainable healthcare top-of-mind**

*GP9 Yes, well, we've mainly just brought it to people's attention again*

#### *11. Environmental context and resources*

**A. Using the natural and digital environment**

##### a. Have a Green GP practice

Participants indicate that having references to sustainability in the GP practice helps them implement sustainable care. There are various ways this is expressed:

1. For example, it helps to have physical reminders of sustainability in the GP practice. These might include a general recycling bin and a separate recycling bin for medications or not having a printer on each desk but rather just one for the entire practice.
2. Elements specifically aimed at informing patients, such as infographics or posters in the waiting room about sustainable care or sustainable care projects ('no risk, no glove') or showing information about sustainable care on the waiting room screen.
3. Highlighting the focus on sustainable care, for example on the website or in a newsletter.

This helps legitimize incorporating sustainable care into consultations.

*GP10 10:35: Well, at my practice, we had a statement on our website explaining that we highly value sustainability—what that means to us and what we're doing about it—and we also displayed a message on the waiting room screen. That way, patients know you're actively working on it, and it also lends more credibility to the fact that you're incorporating it into your consultations. I think that really helps, too.*

##### b. Use the possibilities of GP software

Incorporating green options into GP IT systems makes it easier to provide sustainable care. This includes information on the number of tablets per pack of medication, making it possible to prescribe quantities tailored to the pack size, when medically justified. Information on the $CO_2$ emissions of different medicines would also be helpful

in making more sustainable choices. Several GPs also mention that they use the repeat prescriptions list (prescription buffer); this allows them to check whether unnecessary medication is being requested or whether it is possible to switch a patient to a more sustainable alternative. A specific GP information system also helps by placing the sustainable option at the top when prescribing in accordance with the formulary.
*GP 3 3:5 [When] I see the metered-dose inhaler [...] in the list, I start the conversation about it straight away.*
*GP1 1:25: As I understand it, if you click on 'prescribe according to the formulary', the powder [the powder inhaler] is listed above the metered-dose inhaler. [...] So both are listed, but the first one you can click on is the correct one, and I think that helps a great deal.*

##### c. Use patient information materials

It helps to have devices, questionnaires or information packages, to support more sustainable healthcare. For example, using the lifestyle wheel ('Leefstijlroer') when starting a conversation about lifestyle medicine, or information from a national medical website GPinfo.nl (thuisarts.nl) or nutrition information on the Netherlands Nutrition Centre ('Voedingscentrum') about plant-based foods. One participant stated that it would help her even more if there was a page available on sustainable healthcare on thuisarts.nl, so she could refer her patients to that website.
*GP 9 9:24: After many consultations, I email the patient some additional information from Thuisarts [GPinfo.nl]. [...] So it would be helpful if [sustainable healthcare] would be clearly explained there for the patient and if I could send that to the patient as well.*

##### d. Use supporting devices

Devices such as the In-Check DIAL can contribute to the provision of sustainable healthcare. In case of the In-Check DIAL, it helps patients make the switch from the less sustainable option – the pressurised metered-dose inhaler (pMDI) – to the more sustainable option – the dry powder inhaler (DPI).
*GP12 12:22 [The In-Check DIAL] Yes, that really did help our practice nurse a great deal. [...] To convince people that: 'This [a DPI] is definitely just as good for you. You can just use this, yes.'*

###### B. Being a green organization

Being a green organization helps to deliver more sustainable healthcare. This encompasses trying to be green in everything you do, for example recycling bins, using less examination bank paper, recycling medication syringes, using second-hand furniture.

##### a. Have a supportive organisational culture

It helps to have an organization culture that supports sustainable healthcare. For example, creating a sense of co-ownership of the sustainable mindset, by allocating different sustainability projects to different staff members; and create a safe working environment where you can point things out to one another if something isn't being done in a sustainable way.

##### b. Leverage the strengths and opportunities of colleagues

By drawing on your colleagues' skills or expertise, it is possible to work together to provide sustainable care and, as a result, deliver it more effectively. One example might be a practice nurse who has more time for, and experience with, discussing lifestyle interventions. By referring a patient to that nurse, you are providing sustainable care by making use of a colleague's skills.

##### c. Have organizational sustainability actions §

By carrying out an annual medication review for selected patients, overprescribing issues are routinely identified, making it easier to maintain a critical eye on long-term medication and thus discontinue unnecessary medication. It can also help to visit patients more frequently, thereby preventing medication from being used for too long or causing too many side effects; and thus, ensuring it remains sustainable.

##### C. Have sustainability in guidelines

It helps that sustainability is increasingly incorporated into medical guidelines in several ways. For the Green GPs in our sample, it serves as a back-up when discussing sustainable healthcare with colleagues, as they can say: It's not just me doing this, but it's in the guidelines as well. But also, because, as a doctor, you can't know everything, and it's helpful to know that you don't have to look up what the sustainable option is – it's already set out in the guidelines. It also helps GPs and GPs in training to learn more about sustainable healthcare.

*GP11 11:31 Well, what I particularly like is that you can also justify things to your colleagues, can't you? You can just say, 'Look, it's in the guidelines too.'*

*GP8 8:16 Yes, because you don't know exactly everything about it [when it comes to sustainable healthcare]. Just like with those NSAIDs. The advice on which ones are and aren't safe changes every so often. So, you think, how am I supposed to keep track of that? And then it's nice when you can find that information clearly set out somewhere.*

*GP9 9:18 Look, I don't prescribe COPD or asthma medication for new patients every day, so you do end up referring back to those guidelines quite often – and the same goes for the junior doctors and registrars. Yes, so it's absolutely vital that those guidelines are included. And that any updates are now also assessed through a green lens. No, well, the fact that it's in your reference material – that's absolutely very important.*

##### D. Have supportive legislation

It would also help if there were more support from the legislation. For example, if medication were only reimbursed once non-pharmacological options had been tried for a considerable period of time. This would allow doctors to shield themselves, because you are not the one to blame, but the law is.

##### E. Make use of unexpected events

During the winter of 2024 there was a shortage of pMDI's, this helped Green GPs to start the conversation about switching from a pMDI to a DPI with their patients.

*GP9 9:5: The shortages back then certainly helped, didn't they? The problem with salbutamol – that simply forced you to make that switch. Not so much out of environmental considerations, but simply because it wasn't available. Yes, that does help to get a group to switch.*

##### F. Time

Within your consultations, but also within your workweek it helps to make time for sustainable healthcare. Within your consultations this time helps to better explain the different options, or sustainable healthcare, or to discuss lifestyle changes. During your workweek it helps to specifically allocate time and headspace to sustainable healthcare, for example by picking out one half-day a week to specifically focus on sustainable healthcare. Lastly, it helps to allocate time to sustainable healthcare meetings, for example by planning a sustainable day each month to have someone tell something about sustainable healthcare or having Green Pharmacology meetings.
*GP 1 1:28 That's Sustainable Tuesday and Sustainable Thursday. So once a month, [...] we have a sort of sustainability training.*
It also works the other way around, that you allow yourself not to focus so much on sustainability when you don't have the time and headspace; this helps to sustain the sustainable flow in the long run.
*GP3 3:13: A request for a repeat prescription – you know, when you're so busy and you really don't have any time left, and you think: 'Oh, never mind, I'm not going to ring the patient; I'll just click it through.' It's a bit of a shame, really, but well, that's how it goes sometimes. And then I say to myself, 'I'll just try again next time, then.'*
Lastly, it helps to acknowledge that sustainability may also save you time, as stated in beliefs about consequences, by lowering the amount of actions you have to do.

##### G. Use green working agreements

It helps to establish practical working agreements with other healthcare providers in the local area, making it easier to deliver sustainable care. The key player in this is the pharmacist. An example of this is agreeing that the pharmacist may choose the exact type of powder inhaler based on what is available on the shelf, provided there is no difference for the patient, in order to prevent medication from expiring. Or that the pharmacist may dispense the measured quantity from a pack if the doctor prescribes a one-week supply. Or, for instance, the already legally established limit of dispensing a maximum of two weeks' supply on the first dispensing.

#### *12. Social influences*

##### A. The Green norm

##### a. Have sustainable healthcare as the norm within our GP practice

It helps if sustainability is the norm throughout the GP practice; that makes it easier to follow suit.
*GP11 11:45 We all cycle or take the train [to work]. That was partly why I chose this staff.*
*GP1 1:26 So my fellow GPs all take the same view on this, so they're also quite convinced on this issue [sustainable healthcare]. Generally speaking, they're all relatively sustainable. So, I encounter little resistance. It goes quite smoothly, then.*

*GP6 6:23 If, for example, you look at how they [colleagues.] already sort their waste properly, then it is certainly easier to go along with that.*

##### b. Have sustainable healthcare as the norm for GPs

Participants indicate that adopting a green approach felt already as the norm for general practitioners, as the non-medication option – which is often also the more sustainable option – has always been the first step in GP guidelines. However, they feel that this is now not only the norm among GPs, but that patients also regard the focus on non-medication advice or lifestyle changes as the norm. Legislative support also helps; for example, the law stipulates that pharmacists may only dispense a two-week supply on the first prescription. This makes it easier to choose the more sustainable option.
*GP1 1:3 That it's in the guidelines. [...] If you prescribe it in accordance with the formulary [prescription software, red], it's now listed at the top too, so it's simply standard care – it's not something only I do or believe in. But it's just that this is the standard, and I, personally, really like that.*

##### c. Have sustainable healthcare as the norm for my patients

*GP12 12:24: Yes, and people are used to it too – they've been for quite some time, actually.*

#### d. Have sustainable healthcare as the norm in society

As sustainability in general, and sustainable healthcare in particular, are increasingly receiving media attention, it is becoming a topic that is more widely accepted in society to discuss. This makes it easier to provide sustainable healthcare.
*GP 11 11:42 Yes, so there's more and more information about it out there. People don't find it as strange anymore, you know, so there's less resistance, which makes it easier.*
*GP9 9:7: Due to the shortages and the recent media reports about the pMDI's, you can also see [...] that awareness amongst patients is growing.*
*GP11 11:41: Well, I suppose it's become more the norm, hasn't it? So that does help. Yes, the fact that you're not some sort of odd one out for doing that.*

##### B. Social support

#### a. Have social support in our GP practice §

It helps to have social support in delivering sustainable healthcare in your local GP practice.
*GP4 4:17 And the fact that he [co-owner of the GP practice] supports that [providing sustainable care] does help, of course. I mean, otherwise you're a bit like a voice crying in the wilderness, and that just works against you. [...] Yes, in terms of sustainability, there are things we can introduce, and he's fine with all of that.*

#### b. Have social support from network partners

It helps to have social support from other local partners, such as pharmacists, to deliver more sustainable healthcare.

##### c. Have social support from like-minded GPs

It helps to have the social support of other Green GPs. Within your local GP network, but also on a national level. This makes that you feel less as the odd one out, or a voice crying in the wilderness; and helps to implement sustainable healthcare even more.
*GP5 5:3: I remember that back then, during those first few meetings [...] you really thought: 'Wow, there are others out there working on this too.'*

#### *13. Emotions*

##### A. Enjoy delivering sustainable healthcare

Most participants report having fun or just liking it when engaging in sustainable healthcare, which encourages them to keep going.

##### B. Reframe negative experiences into motivation to try harder ◊

Negative emotions, like the feeling you have dropped the ball in terms of sustainable healthcare, actually are a motivation for some participants, to try even harder next time to make the sustainable healthcare option work.

#### *14. Behavioural regulation*

##### A. Use self-monitoring

It helps to actively seek feedback on whether providing sustainable care is effective for the patient. This can be done, for example, by following up with a patient and making any necessary adjustments. This helps you learn what does and does not work well in the context of sustainable care.

##### B. Prepare for sustainability conversations

Having standard phrases for a conversation about sustainability makes it easier to provide sustainable care. This can be done, for example, by asking the patient for permission to explain about sustainable care, or by focusing the conversation not on the benefits of the sustainable option, but on the downsides of the medication-based option (such as side effects).

##### C. Pick your battles wisely

It helps to ensure that you are able to provide sustainable care in the long term by making specific choices about when to opt for the sustainable option and when not to, if doing so requires more energy or time at that particular moment.
*GP4 4:1 It also depends a bit on the day you're having. Whether you're feeling on top of things, so to speak – just as you'd consider whether to prescribe antibiotics for someone coughing sitting in front of you, and you can say with complete conviction, 'Well, that's not necessary at all.' You might be better at that on some days than others.*

##### D. Make sustainable healthcare a habit

It helps when the sustainable option becomes your own habit. This makes it easier to deliver sustainable healthcare.

*GP12 12:5 So, certainly when it comes to first-time prescriptions, a powder inhaler is actually prescribed fairly standardly – by practically everyone in our practice*.

##### E. Make it a project

By making it a project and taking small steps to achieve that, it is easier to deliver sustainable healthcare. For example, starting to ring patients with the non-sustainable inhalation medication and proposing them to consider switching.

#### 7. Figure S1

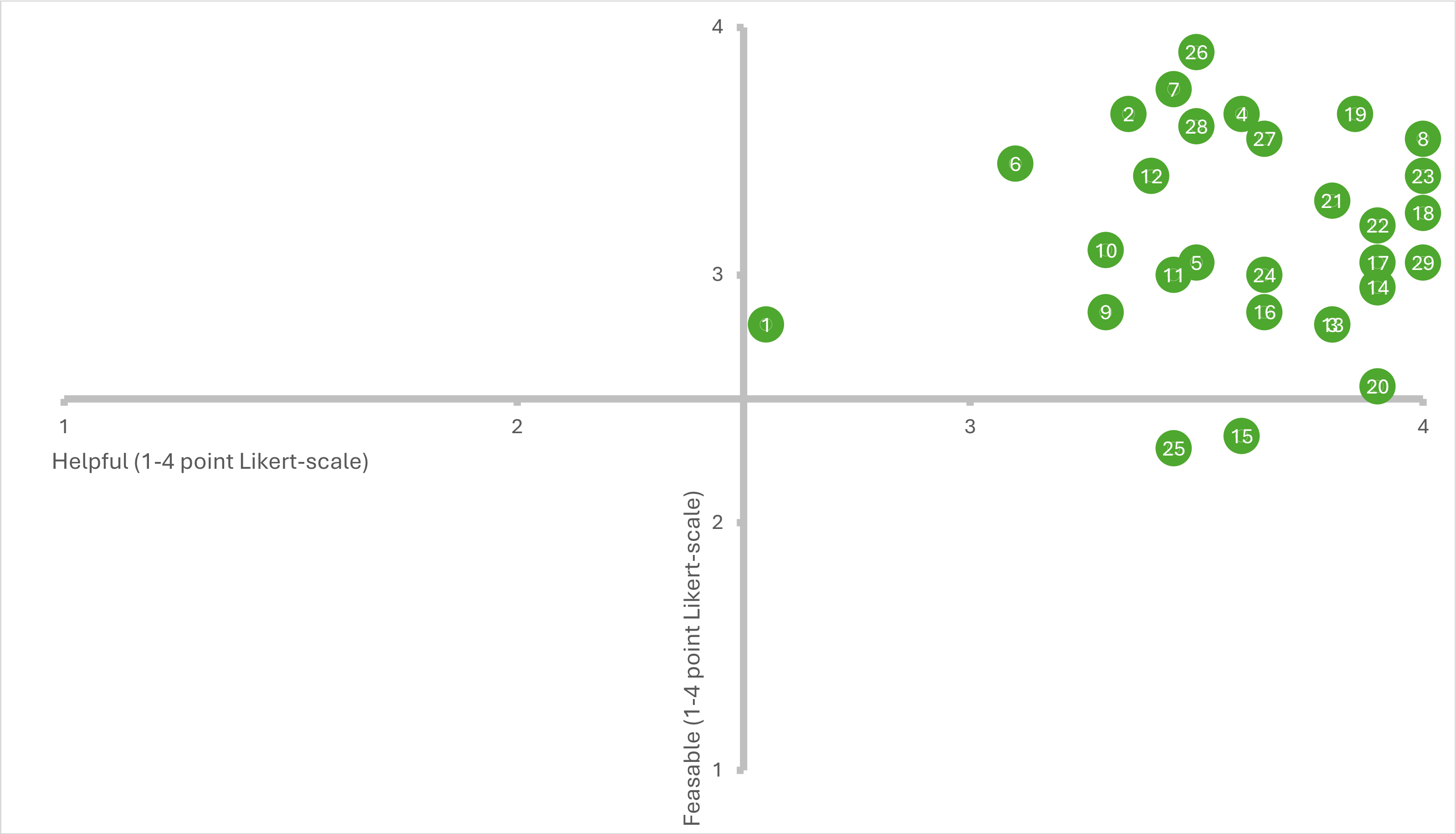

*Figure S1 – Prioritization of actionable facilitators for behaviour change for GPs from environmentally less sustainable to environmentally more sustainable. Numbers per facilitator are in the legend. Helpful on x-axis and Feasible on y-axis, both on 1-4 Likert scale.*

Legend

To deliver more sustainable healthcare it helps to:

1. Approach every patient as a new opportunity to raise the issue of sustainability
2. Decide when to apply sustainable care and when not to
3. Maintain good cooperation with the pharmacy
4. Communicate about sustainability in the practice through posters, on-screen presentations in the waiting room, recycling bins, etc.
5. Schedule follow-up consultations to review medication in order to prevent medication wastage and chronic use where possible
6. Experience every instance of not prescribing, prescribing sustainably or taking another sustainable care action as a success
7. Recognise the additional benefits of sustainable care for me as a doctor
8. Make sustainability and prevention part of my professional identity
9. Highlight the co-benefits of the sustainable option to patients
10. Prioritise the patient’s health within sustainable care
11. Giving and receiving positive feedback on sustainable care from colleagues
12. Convey confidence to the patient when communicating about sustainable care
13. Approach sustainable care as projects
14. Have a standardised sustainable recommendation or guideline across the practice or region
15. Gain knowledge about the $CO_2$ emissions of certain medications; and being able to make comparisons for colleagues or patients
16. Acquire skills to communicate about sustainable healthcare
17. Pay special attention to sustainable healthcare within the team
18. Be able to refer to sustainable care in the GP guidelines and on GPinfo.nl as a resource when communicating with patients and colleagues
19. Incorporate sustainable care into my routine / make it my default option
20. Know how many tablets are in a pack of medication, so as to be able to prescribe in appropriate quantities
21. Be aware that other GPs are working on green healthcare at local, regional and national levels
22. Be able to provide scientific evidence for sustainable healthcare
23. Have knowledge of sustainable healthcare, including a healthy lifestyle
24. Critically question my routines as a GP
25. Recognise non-sustainable medication options and use this as an incentive to switch to more sustainable, equivalent alternatives
26. Start with simple sustainable options in the consultation room
27. Take small steps with the patient towards more sustainable care, rather than trying to make everything sustainable all at once
28. Try out sustainable care options first in simple consultations
29. Use tools to support a healthier lifestyle and sustainable care for patients
